# Cardiac Microstructural Abnormalities Identify Women at Risk of Incident Heart Failure

**DOI:** 10.1101/2020.01.03.20016477

**Authors:** Alan C. Kwan, Emmanuella Demosthenes, Trevor Nguyen, Eric Luong, Gerran Salto, Ewa Osypiuk, Plamen Stantchev, Elizabeth H. Kim, Pranoti Hiremath, Vanessa Xanthakis, Ramachandran S. Vasan, Susan Cheng

## Abstract

**Background:** Incidence of congestive heart failure is difficult to predict by standard methods. We have developed a method called the signal intensity coefficient that uses echocardiographic texture analysis to quantify microstructural changes which may occur in at-risk patients prior to development of a clinical heart failure syndrome.

**Methods:** Participants from the Framingham Offspring Cohort study who attended the 8^th^ visit and received screening echocardiography were included. Participants were followed for a mean of 7.4 years for incident congestive heart failure. Cox proportional hazards modeling was used to assess the hazard ratio of signal intensity coefficient in the top quartile of values versus other quartiles in the total and sex-stratified population.

**Results:** 2511 participants with interpretable echocardiography and no history of congestive heart failure, stroke, or myocardial infarction were included in this study. The top quartile signal intensity coefficient had a hazard ratio of 1.83 (p=0.0048) for incident heart failure. When additional clinical risk factors were added to the model, this became non-significant. Within women, an elevated hazard ratio was significant in multiple models including age and hypertensive medication use. Models were not significant in men.

**Conclusions:** Elevated signal intensity coefficient is associated with an increased risk of incident congestive heart failure. This trend remains significant in women after inclusion of age and hypertensive medication use. The signal intensity coefficient may be able to identify patients at risk of developing congestive heart failure using echocardiographic texture analysis.

## INTRODUCTION

Heart failure affects 6.2 million Americans over 20 years of age and is expected to increase by 46% from 2012 to 2030.^1^ While management of heart failure after diagnosis has improved through modern therapies, our ability to predict heart failure remains modest at best and we lack specific markers to identify patients at risk.^2,3^ The Signal Intensity Coefficient (SIC) is an echocardiographic marker of tissue microstructure, correlating with interstitial fibrosis and microfiber disarray.^4,5^ It has been utilized to identify preclinical changes in the myocardium of patients with hypertension, hypertrophic cardiomyopathy gene carriers, and obese patients.^4,6,7^ We hypothesized that the SIC is able to identify preclinical changes in patients who subsequently develop incident heart failure in the Framingham Offspring Cohort. By identifying the risk of future heart failure in patients prior to clinical disease, we may be able to more effectively prevent heart failure.

## METHODS

### Study Sample

The study sample was drawn from the Framingham Offspring study, which has been previously described.^8^ Participants received routine examinations approximately every 4 years. Those who attended the 8^th^ examination between 2005-2008 were eligible for inclusion in this study. The study protocols were approved by the IRB and all participants provided written informed consent.

### Clinical Assessment

Screening echocardiography and clinical and serological assessment of cardiometabolic risk factors was performed during the participants’ routinely scheduled eighth examination. Participants with adequate quality echocardiography for texture analysis and without pre-existing history of heart failure, stroke, or myocardial infarction were included (Figure 1). Participants were monitored longitudinally as per standard protocol, with scheduled follow-up at visit 9 between 2011-2014 and interim data collection via medical record review. The primary outcome was incident heart failure. Diagnosis of heart failure was made using standard Framingham criteria.^9^ Heart failure was defined as the presence of two major, or one major and two minor criteria. Major criteria include paroxysmal nocturnal dyspnea or orthopnea, distended neck veins, rales, radiographic cardiomegaly, pulmonary edema, S3, hepatojugular reflux, and weight loss on diuretic therapy; minor criteria include bilateral ankle edema, night cough, dyspnea on exertion, hepatomegaly, pleural effusion, radiographic pulmonary vascular redistribution, decrease in vital capacity, and tachycardia. All new diagnoses were adjudicated by a panel of three experienced physicians conducting medical record review.

**Figure 1.**
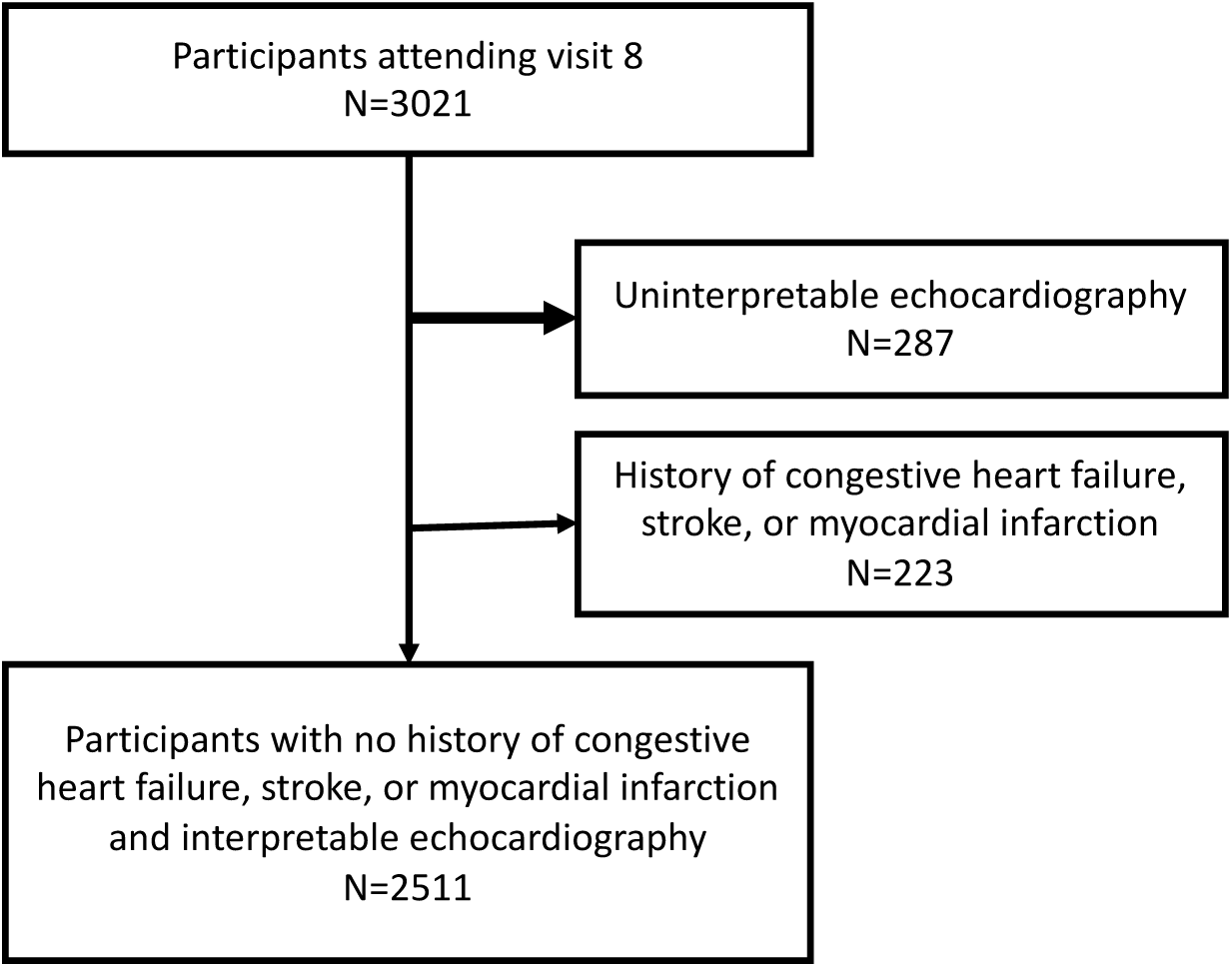
Flow diagram for patient inclusion.

### Echocardiographic Assessment and Analysis

Transthoracic echocardiography was obtained using standard techniques, as previously described.^10,11^ Echocardiographic measurement was performed per Framingham Offspring Study standard.^12^ Quantitative tissue analysis was performed as previously described.^4-7^ Briefly, transthoracic B-mode parasternal long axis views were selected at end diastole. Images were inspected for quality and ability to visualize the mid-to-basal inferiolateral myocardial/pericardial interface. Static images (8-bit DICOM or JPG) were analyzed using the ImageJ software platform (v1.46, National Institutes of Health, Bethesda, MD, USA). A macro was applied to a user-specified region of interest drawn at the myocardial/pericardial interface, which provided a hierarchical distribution of signal intensities (0-255) of the pericardium next to the mid-to-basal inferiolateral myocardial wall. SIC was calculated as 1-(p/256), where p is the 25^th^ percentile signal intensity within the pericardial region of interest. Myocardial structural index (MSI) was defined as SIC/0.13 + RWT/0.05 (i.e. the sum of the standardized value of SIC and the standardized value of RWT derived from a healthy reference subsample. Both SIC and MSI were divided into quartiles, with the primary exposure being participants in the top quartile of SIC (which we will refer to as “high SIC”) or MSI (“high MSI”) versus the other three quartiles.

### Statistical Analysis

Sample characteristics were displayed as mean and standard deviation for normally distributed variables, median and interquartile range for non-normal variables, and as a percent for categorical variables. Female versus male baseline characteristics were compared using the student’s T test for normally distributed continuous variables, Wilcoxon test for non-normally distributed continuous variables, and chi-squared test for categorical variables. Logistic regression was performed to test association between clinical risk factors of age, sex, body mass index (BMI), total cholesterol/high density lipoprotein, systolic blood pressure, hypertensive treatment, diabetes, and smoking with high SIC in the overall population as well as stratified by sex. Multivariable-adjusted Cox proportional hazards models were used to examine the relationship between high SIC and high MSI with the incidence of new-onset HF in sex-pooled and sex-specific analyses. These were performed with and without adjustment for conventional risk factors. Age was stratified into quartiles for Cox analysis as the continuous variable failed the proportionality assumption. For the overall analysis, model 1 included high SIC, age, and sex as a basic model. Model 2 was equivalent to model 1 plus hypertensive medication use, due to known correlation of SIC with history of hypertension. Model 3 was model 2 plus standard Framingham risk factors of BMI, systolic blood pressure, diabetes, smoking, and total cholesterol to HDL ratio. The same models were retested in male and female populations, with sex removed as a variable. Secondary analyses included testing with high MSI instead of high SIC. Sex/high SIC and sex/high MSI interaction terms were tested in total population models as well. Analyses were performed using R (v 3.6.1) with Rstudio (1.2.5019). Alpha was prespecified as 0.05.

## RESULTS

The study population consisted of 2511 participants, which were majority female (56%) (Table 1). Mean age was 65 years old, and participants were followed for a mean of 7.4 years. Mean BMI in both men and women was 28.1, classified as “overweight”. 39% of participants were using lipid-lowering medications, 45% were on antihypertensive medications, and 7.5 percent were on diabetes medications. The mean LVEF was normal at 67.5%. Statistically significant differences were noted between all clinical characteristics of men and women except for age and smoking rate; however, most differences were small. Mean SIC in the total population was 36.4±14.6%, with 39.8±15.1% in men and 33.8±13.6% in women, a significant difference (p<0.0001). The mean MSI was 8.56±1.47 in the total population, 8.98±1.49 in men, and 8.2±1.38 in women (p<0.0001).

**Table 1.** Characteristics of participants for total population and by sex.

|  | **Total** | **Women** | **Men** | **p** |
| --- | --- | --- | --- | --- |
| N | 2511 | 1414 (56%) | 1097 (43%) | <0.0001 |
| Age (y) | 65.7±8.8 | 65.8±8.7 | 65.5±8.8 | 0.39 |
| Follow-up Time (y post-echo) | 7.4±1.7 | 7.5±1.6 | 7.2±1.9 | <0.0001 |
| BMI (kg/m<sup>2</sup>) | 28.1±5.3 | 27.6±5.8 | 28.7±4.5 | <0.0001 |
| Total Cholesterol/HDL | 3.47±1.05 | 3.27±0.95 | 3.74±1.11 | <0.0001 |
| Lipid-Lowering Medication Use (%) | 981 (39%) | 498 (35%) | 485 (44%) | <0.0001 |
| SBP (mmHg) | 128±17.1 | 128±17.4 | 129±16.6 | 0.0111 |
| DBP (mmHg) | 73.9±9.94 | 72.6±9.68 | 75.6±9.99 | <0.0001 |
| Antihypertensive Medication Use (%) | 1122 (45%) | 583 (41%) | 539 (49%) | <0.0001 |
| Diabetes (%) | 289 (12%) | 121 (8.6%) | 168 (15%) | <0.0001 |
| Fasting Glucose (mg/dl) | 106±22.2 | 102±18.8 | 110±25.4 | <0.0001 |
| Diabetes Medication Use (%) | 189 (7.5%) | 89 (6.3%) | 100 (9.1%) | <0.01 |
| Smoker (%) | 217 (8.6%) | 135 (9.5%) | 82 (7.5%) | 0.08 |
| eGFR (mL/min/1.73m<sup>2</sup>) | 81.2±18.2 | 79.7±18.5 | 83.1±17.5 | <0.0001 |
| LVEF (%) | 67.5±6.86 | 68.9±6.43 | 65.7±6.96 | <0.0001 |
| LVMI (%) | 88.4±19.8 | 81.0±16.0 | 98.0±20.0 | <0.0001 |

BMI: Body Mass Index, SBP: Systolic Blood Pressure, DBP: Diastolic Blood pressure, eGFR: estimated Glomerular Filtration Rate, LVEF: Left Ventricular Ejection Fraction, LVMI: Left Ventricular Mass Index.

High SIC was associated with multiple risk factors in the total population by logistic regression, including higher age, male sex, higher BMI, higher total cholesterol to HDL ratio, use of hypertensive medication therapy, and presence of diabetes (Table 2). In women, age, BMI, total cholesterol to HDL ratio, systolic blood pressure, hypertensive therapy, and diabetes were significant. Systolic blood pressure had not previously been significant in the total population. In men, similar factors were significant with the exception of systolic blood pressure (Table 2).

**Table 2.** Logistic Regression for clinical risk factors associated with high signal intensity coefficient.

| Total Population | $\beta$ | 95% CI | SE | p | Women $\beta$ | 95% CI | SE | p | Men $\beta$ | 95% CI | SE | p |
| --- | --- | --- | --- | --- | --- | --- | --- | --- | --- | --- | --- | --- |
| Sex* | -0.77 | (-0.96,-0.59) | 0.09 | <0.0001 |  |  |  |  |  |  |  |  |
| Age ** | 0.03 | (0.02,0.04) | 0.01 | <0.0001 | 0.04 | (0.02,0.05) | 0.01 | <0.0001 | 0.03 | (0.01, 0.04) | 0.01 | <0.0001 |
| BMI | 0.12 | (0.10,0.14) | 0.01 | <0.0001 | 0.12 | (0.10,0.15) | 0.01 | <0.0001 | 0.12 | (0.09,0.15) | 0.02 | <0.0001 |
| Total Cholesterol/HDL | 0.24 | (0.16,0.33) | 0.04 | <0.0001 | 0.36 | (0.22,0.49) | 0.07 | <0.0001 | 0.16 | (0.04,0.27) | 0.06 | 0.01 |
| SBP | 0.00 | (0.00,0.01) | 0.00 | 0.11 | 0.01 | (0.00,0.02) | 0.00 | 0.00 | 0.00 | (-0.01,0.01) | 0.00 | 0.48 |
| HTN treatment | 0.38 | (0.19,0.57) | 0.10 | 0.00 | 0.49 | (0.21, 0.77) | 0.14 | 0.00 | 0.27 | (0.01,0.54) | 0.13 | 0.04 |
| Diabetes | 0.60 | (0.33,0.86) | 0.13 | <0.0001 | 0.79 | (0.37,0.1.19) | 0.21 | 0.00 | 0.47 | (0.13,0.81) | 0.17 | 0.01 |
| Smoking | -0.04 | (-0.40,0.30) | 0.18 | 0.82 | 0.07 | (-0.42,0.53) | 0.24 | 0.76 | -0.17 | (-0.70,0.22) | 0.26 | 0.52 |

BMI: Body Mass Index, HDL: High density lipoprotein, SBP: Systolic Blood pressure, HTN: Hypertension.
All measures were adjusted for age and sex unless otherwise noted.
\* Adjusted by age only
\*\* Adjusted by sex only

High SIC alone for incident congestive heart failure had a hazard ratio of 1.8262 (p=0.0048) in the sex-pooled total population. Inclusion of other risk factors in any of the three models resulted in a non-significant hazard ratio within the total population (Table 3). However, analysis with the inclusion of sex by high SIC interaction term showed significant change in model 1 (Supplemental Table 1), justifying further sex-stratified analyses. Within women, mean SIC was 28.3% in the non-high SIC category and 56.4% in the high SIC category. In men, mean SIC was 30.7% in the non-high SIC category and 57.8% in the high SIC category. In Cox models for women, high SIC was significant in both Model 1 and Model 2, and subsequently not significant in Model 3 (Table 3). In model 1, the hazard ratio associated with high SIC was 132% higher for those with high SIC versus not high SIC. In model 2, the hazard ratio was 98% higher for congestive heart failure with high SIC. Within men, high SIC was not significant in any of the three models. Analysis with high MSI was performed and substituted for high SIC (Table 4). In the total population, Model 1 showed a 69% increase in hazard ratio for congestive heart failure with high MSI. The analyses in model 2 and 3, as well as those for the sex-stratified analyses, were not significant. Kaplan Meier analyses were performed for participants with high SIC versus non-high SIC in both the total population (Figure 2A) and the sex-stratified population (Figure 2B). High SIC was associated with increased incidence of congestive heart failure in the total population, as well as for women in the sex stratified population, but not for men.

**Table 3.** Hazard ratio for high signal intensity coefficient, cox proportional hazards models.

| Sex Stratified | Women |  |  |  | Men |  |  |  | Total Population |  |  |  |
| --- | --- | --- | --- | --- | --- | --- | --- | --- | --- | --- | --- | --- |
|  | Hazard Ratio | 95% CI | SE | p | Hazard Ratio2 | 95% CI | SE2 | p | Hazard Ratio | 95% CI | SE | p |
| Model 1 | 2.32 | (1.20,4.50) | 0.34 | 0.01 | 0.98 | (0.57,1.70) | 0.28 | 0.95 | 1.36 | (0.89,2.08) | 0.22 | 0.16 |
| Model 2 | 1.98 | (1.02,3.84) | 0.34 | 0.04 | 0.99 | (0.57,1.72) | 0.28 | 0.98 | 1.32 | (0.86,2.02) | 0.22 | 0.20 |
| Model 3 | 1.69 | (0.84,3.41) | 0.36 | 0.14 | 0.78 | (0.44,1.37) | 0.29 | 0.39 | 1.04 | (0.67,1.62) | 0.23 | 0.86 |

Model 1: High signal intensity coefficient and sex stratified by quartiles of age. Model 2: Model 1 + hypertensive medication use. Model 3: Model 2 + body mass index, systolic blood pressure, diabetes, smoking, and total cholesterol to high density lipoprotein ratio. SE: Standard Error.

**Table 4.** Hazard ratio for high myocardial structural index, cox proportional hazards models.

| Sex Stratified | Women |  |  |  | Men |  |  |  | Total Population |  |  |  |
| --- | --- | --- | --- | --- | --- | --- | --- | --- | --- | --- | --- | --- |
|  | Hazard Ratio | 95% CI | SE | p | Hazard Ratio | 95% CI | SE | p | Hazard Ratio | 95% CI | SE | p |
| Model 1 | 1.59 | (0.74,3.42) | 0.39 | 0.23 | 1.70 | (0.97,2.96) | 0.28 | 0.06 | 1.69 | (1.08,2.64) | 0.23 | 0.02 |
| Model 2 | 1.25 | (0.58, 2.70) | 0.39 | 0.57 | 1.59 | (0.90,2.78) | 0.29 | 0.11 | 1.52 | (0.97,2.39) | 0.23 | 0.07 |
| Model 3 | 1.10 | (0.50,2.42) | 0.40 | 0.81 | 1.40 | (0.79,2.50) | 0.29 | 0.25 | 1.31 | (0.83,2.08) | 0.24 | 0.25 |

Model 1: Myocardial structural index and sex stratified by quartiles of age. Model 2: Model 1 + hypertensive medication use. Model 3: Model 2 + body mass index, systolic blood pressure, diabetes, smoking, and total cholesterol to high density lipoprotein ratio. SE: Standard Error.

**Figure 2.**
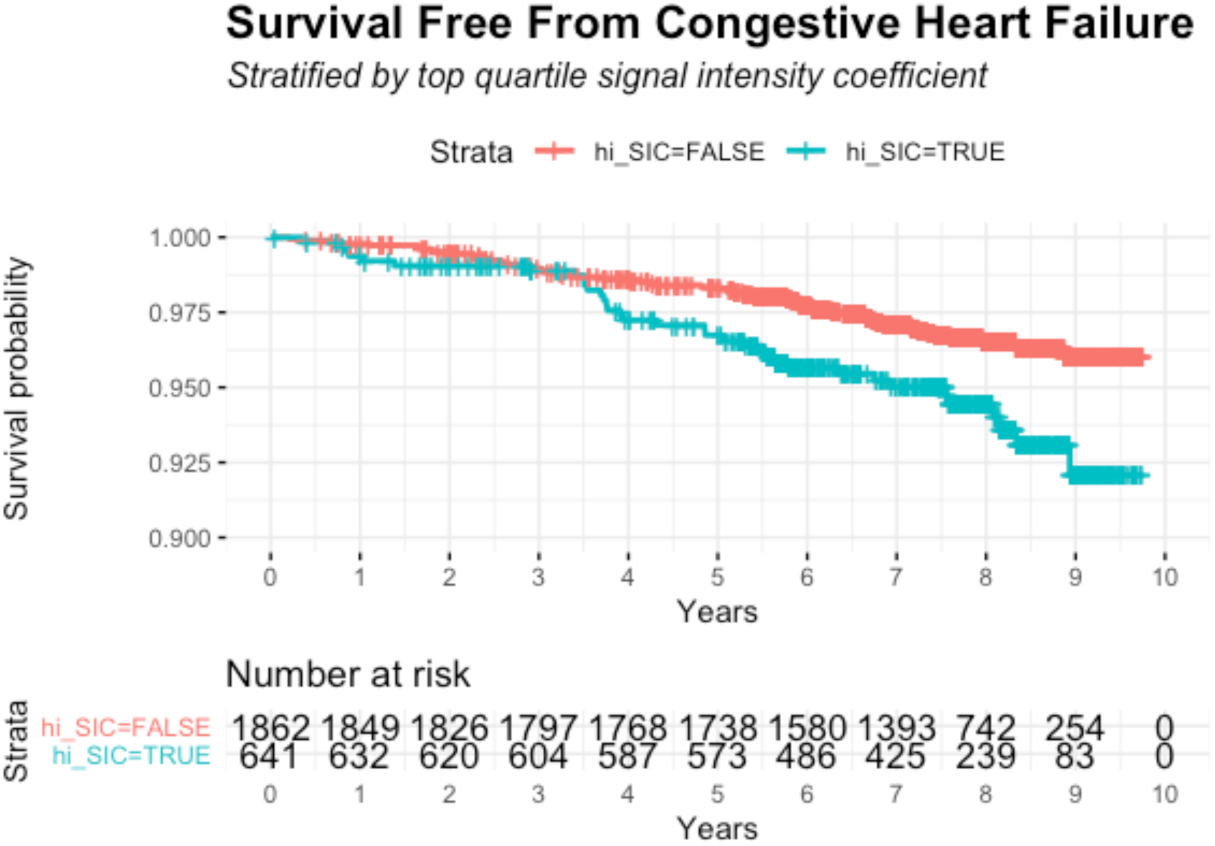
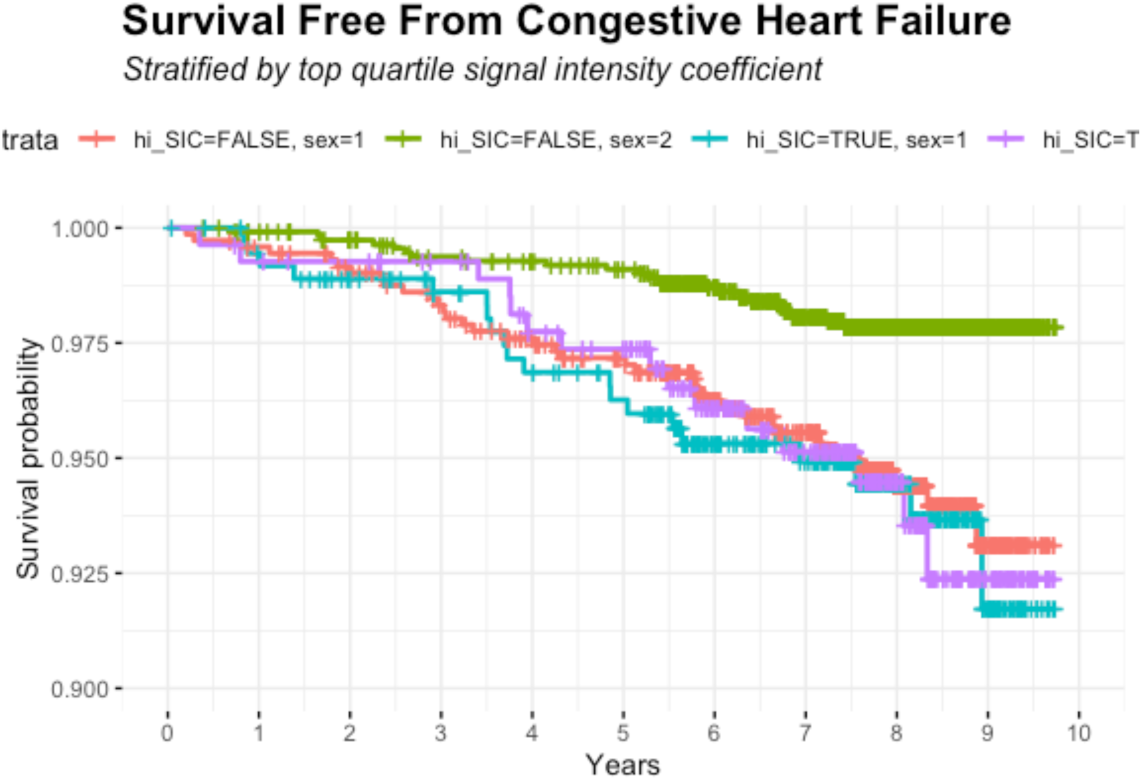
A) Kaplan Meier Curve for Survival Free from Congestive Heart Failure Stratified by Signal Intensity Coefficient. Red: participants with Signal Intensity Coefficient in lower three quartiles. Blue: participants with Signal Intensity Coefficient in top quartile **B) Kaplan Meier Curve for Survival Free from Congestive Heart Failure Stratified by Signal Intensity Coefficient and Sex**. Red: male participants with Signal Intensity Coefficient in lower three quartiles. Blue: male participants with Signal Intensity Coefficient in top quartile. Green: female participants with Signal Intensity Coefficient in lower three quartiles Purple: female participants with Signal Intensity Coefficient in top quartile.

## DISCUSSION

Microstructural abnormalities are detectable by the SIC prior to clinically evident changes in patients with hypertension^4^ as well as asymptomatic hypertrophic cardiomyopathy carriers.^6^ In a large longitudinal cohort of participants followed for incident heart failure, we found that SIC in the top quartile of participants have a significantly higher risk for incident heart failure, though this effect is attenuated by inclusion of other clinical risk factors. In this study analyzing the association of elevated SIC with incident heart failure, the findings were three-fold. First, while the hazard ratio for high SIC alone was significant (Figure 2A), inclusion of traditional risk factors by Models 1-3 resulted in a non-significant hazard ratio. Differential sex effects were likely, given the significant interaction between sex and high SIC. Secondly, within the sex analyses, we observe that women without high SIC seem to have a different level of risk from women with high SIC, as well as men regardless of SIC category (Figure 2B). Finally, for MSI analyses, this measure seemed to account for sex-related differences, as the total population models were significant, whereas sex stratified models were not.

Incidence of congestive heart failure currently remains difficult to predict. The success of the SIC and MSI in identifying patients at risk of incident heart failure may indicate that pre-clinical microstructural changes are observable prior to the development of clinical symptoms. For the most part, in both the total and sex-stratified population, incidence of heart failure appears to diverge in those with high SIC between 3 and 4 years of follow up. This may help us identify a clinical population that would benefit from early medical intervention to prevent the clinical appearance of congestive heart failure to improve management of these patients. The significant effect in women is particularly notable, as heart failure with preserved ejection fraction is more common in women and lacks effective medical therapies at this time. Therefore, preventative measures may be the most effective way to manage this growing medical issue.

The primary findings of SIC being predictive of incident heart failure are consistent with the hypothesis that microstructural changes occur within the myocardial tissue prior to clinical presentation of congestive heart failure. These changes can be visualized through the SIC and the MSI up to three years prior to clinical diagnosis. However, there are notable differences between men and women. There is a noted higher prevalence of medical therapy for hypertension, dyslipidemia, and diabetes for the men in the population. This may indicate a longer exposure duration to chronic medical conditions within the male participants. On the other hand, we recognize that men *without* high SIC and women *with* high SIC still have a large difference in their SIC value despite similar heart failure incidence, which suggests true differences between these groups. Further exploration of why the SIC is more successful in predicting heart failure in women is required.

While our study has significant strengths as a prospective cohort with a long duration of follow up, limitations are also present. The systematic follow-up of a longitudinal cohort over time can result in significant survivor biases between different populations, as different groups become older. This may be measurable by variables such as sex, or may not be readily measurable for issues such as socioeconomic status or other environmental exposures. Certainly, there is a bias towards patients who are readily able to follow-up and receive echocardiography within this population. The SIC has been validated across settings of gain and platforms but remains a relatively new measure. While our measure attempts to limit reproducibility issues between acquisition methods, it is not feasible to be able to completely control for all potential variables in acquisition. Finally, the criteria for diagnosis of incident heart failure were developed prior to use of many modern markers, such as naturietic peptide levels or echocardiographic tissue doppler. These historical definitions may lack sensitivity for congestive heart failure or differential sensitivity for forms of heart failure (e.g. preserved ejection fraction versus reduced ejection fraction) which may bias our analyses.

In conclusion, the SIC appears to be associated with incident heart failure partially in women. The MSI may be more predictive in the total population than SIC. We believe that these findings suggest that the SIC and MSI may be able to be used to identify at-risk microstructural abnormalities prior to clinical congestive heart failure. It remains to be seen whether or not the SIC is a modifiable risk factor, and prospective studies are required to determine if therapeutic intervention in the high SIC population is able to improve future outcomes.

## Data Availability

Framingham Offspring Cohort Data are publicly available thorough dbGaP

https://www.ncbi.nlm.nih.gov/gap/

---

**Supplementary Table 1.**
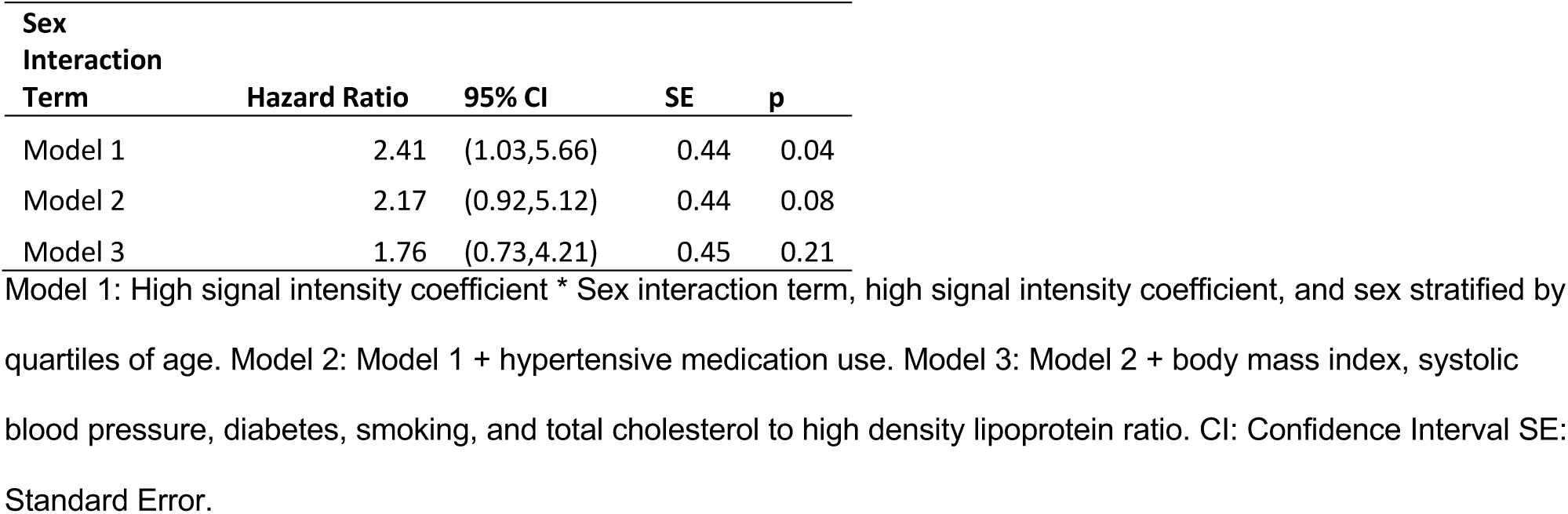
Hazard ratio for sex interaction term added to previous models

